# Homologous Recombination Inquiry Through Ovarian Malignancy Investigations: The Japanese Gynecologic Oncology Group study JGOG3025

**DOI:** 10.1101/2022.07.06.22277241

**Authors:** Kosuke Yoshihara, Tsukasa Baba, Muneaki Shimada, Koji Nishino, Masayuki Sekine, Shiro Takamatsu, Noriomi Matsumura, Hiroshi Yoshida, Hiroaki Kajiyama, Tatsuo Kagimura, Katsutoshi Oda, Yuko Sasajima, Aikou Okamoto, Toru Sugiyama, Takayuki Enomoto

**Affiliations:** Department of Obstetrics and Gynecology, Niigata University Graduate School of Medical and Dental Sciences, Niigata, Japan; Department of Obstetrics and Gynecology, Iwate Medical University, Morioka, Japan; Department of Gynecology, Tohoku University Graduate School of Medicine, Sendai, Japan; Department of Gynecology and Obstetrics, Graduate School of Medicine, Kyoto University, Kyoto, Japan; Department of Obstetrics and Gynecology, Kindai University Faculty of Medicine, Osaka, Japan; Department of Obstetrics and Gynecology, Yokohama Municipal Citizen’s Hospital, Yokohama, Japan; Department of Obstetrics and Gynecology, Nagoya University Graduate School of Medicine, Nagoya, Japan; Translational Research Center for Medical Innovation, Kobe, Japan; Division of Integrative Genomics, University of Tokyo, Tokyo, Japan; Department of Pathology, Teikyo University Hospital, Tokyo, Japan; Department of Obstetrics and Gynecology, The Jikei University School of Medicine, Tokyo, Japan; Department of Obstetrics and Gynecology, St. Mary’s Hospital, Fukuoka, Japan; Department of Obstetrics and Gynecology, Tokai University Graduate School of Medicine, Isehara, Japan; Niigata University Graduate School of Medical and Dental Sciences, Niigata, Japan

**Keywords:** Ovarian cancer, Homologous recombination deficiency, Target-gene sequencing, HITOMI study

## Abstract

The Cancer Genome Atlas has clarified that about 50% of high-grade serous ovarian cancer shows homologous recombination deficiency (HRD). However, the frequency of HRD in Japanese patients with ovarian cancer remains unclear. The aim of JGOG3025 study (NCT03159572) is to identify the frequency of HRD in Japanese patients with ovarian cancer. The JGOG3025 study is a multicenter collaborative prospective observational study involving 65 study sites throughout Japan. We recruited 996 patients who were clinically diagnosed with ovarian cancer before surgery from March 2017 to March 2019, and 701 patients were eligible for JGOG3025 criteria. We used frozen tumor tissues to extract DNA and performed targeted sequencing for 51 genes most of which are HR-associated genes in 701 ovarian cancers (298 high-grade serous, 189 clear cell, 135 endometrioid, 12 mucinous, 3 low-grade serous, and 64 others). HRD was defined as positive when at least one HR-associated gene was mutated. The frequencies of HRD and tumor *BRCA1/2* mutation were 45.2% (317/701) and 18.5% (130/701) in the full analysis set (FAS), respectively. Next, we performed multivariate Cox proportional hazards regression analysis for progression-free survival (PFS) and overall survival (OS). Advanced-stage ovarian cancer patients with HRD had an adjusted hazard ratio of 0.72 (95%CI, 0.55-0.93) and 0.58 (95%CI, 0.39-0.87) for PFS and OS compared to those without HRD (p = 0.013 and 0.009). Our study demonstrated that mutations in HR-associated genes might be associated with their prognosis. Further study will be needed to investigate prognostic impact of each HR-associated gene in ovarian cancer.

## 1. Introduction

Homologous recombination deficiency (HRD) has attracted attention as a new molecular biomarker in gynecologic oncology field^1^. Homologous recombination (HR) repair is one of the repair mechanisms for DNA double-strand breaks, in which the ends of double-strand breaks are precisely repaired^2^. In humans, several genes including *BRCA1/2* and *RAD51* are involved in HR repair, and genomic alterations in these HR-associated genes result in HRD. In HRD cells, DNA repair mechanisms other than HR repair are important, especially poly (ADP-ribose) polymerase (PARP) is focused on a new therapeutic target. PARP inhibition in HRD cells leads to genomic instability and cell death (synthetic lethality)^3^. At present, two PARP inhibitors (olaparib and niraparib) have become clinically available for ovarian cancer in Japan. Interestingly, two phase III trials have demonstrated that niraparib significantly prolonged progression-free survival (PFS) in not only patients with *BRCA1/2* mutations but also those with HRD compared to placebo^4, 5^. PAOLA-1 regimen (olaparib + bevacizumab) also provides a significant PFS benefit in patients with HRD^6^. Thus, HRD status is an important factor in the treatment of ovarian cancer.

The Cancer Genome Atlas (TCGA), a national cancer genome project of the United States, found that about half of high-grade serous ovarian cancers have genomic or epigenomic alterations in the HR-associated pathway such as *BRCA1/2* mutations, *BRCA1* methylation, *CDK12* mutations, and *RAD51C* promoter methylation^7, 8^. However, the frequency of HR-associated mutations in ovarian cancer in Japan remains unclear, especially in non-serous histologic type. Because there are differences in the distribution of histologic types in ovarian cancer between the United States and Japan, the results of TCGA cannot be directly applied to Japan. In addition, clinical significance of HRD remains unclear compared to that of germline *BRCA1/2* mutation. Therefore, it is an urgent issue to investigate the frequency and clinical significance of HRD in Japanese patients with ovarian cancer.

So, on 2017, we started JGOG3025 trial, which was a multicenter collaborative prospective observational study, to clarify the frequency of HRD in Japanese patients with ovarian cancer. We defined the presence of at least one HR-associated gene mutation as HRD in this study. We closed enrollment of the participants at the end of March 2019, and fixed data after 30 months of follow-up. We report the results of final analysis in the JGOG3025 trial.

## 2. Materials and Methods

### 2.1 Patients and study design

A multicenter collaborative prospective observational study (ClinicalTrials.gov, NCT03159572; UMIN 000026303) was conducted in Japan between January 2017 and September 2021. We enrolled the participants for 27 months between January 2017 and March 2019 and the follow-up period was 30 months after the end of enrollment. The target sample size was 700. Study schema is shown in Figure 1. Patients diagnosed with ovarian cancer clinically were enrolled in 65 Japanese Gynecologic Oncology Group (JGOG) institutions throughout Japan (Table S1). After written informed consent for HRD test and biobanking, we obtained tumor tissue samples during surgery. Tumor tissue samples were sent to JGOG-Tohoku Medical Megabank organization (ToMMo) Biobank under controlled temperature. We performed HRD test after the end of enrollment. When patients had the information of germline *BRCA1/2* mutation and allowed us to use this information in this study, germline *BRCA*1/2 mutation status was also submitted to data center. Treatment selection was basically performed according to the Japan Society of Gynecologic Oncology guidelines for the treatment of ovarian cancer, fallopian tube cancer, and primary peritoneal cancer^9, 10^. All evaluations were scheduled according to the clinical practice standards of each institution.

**Figure 1.**
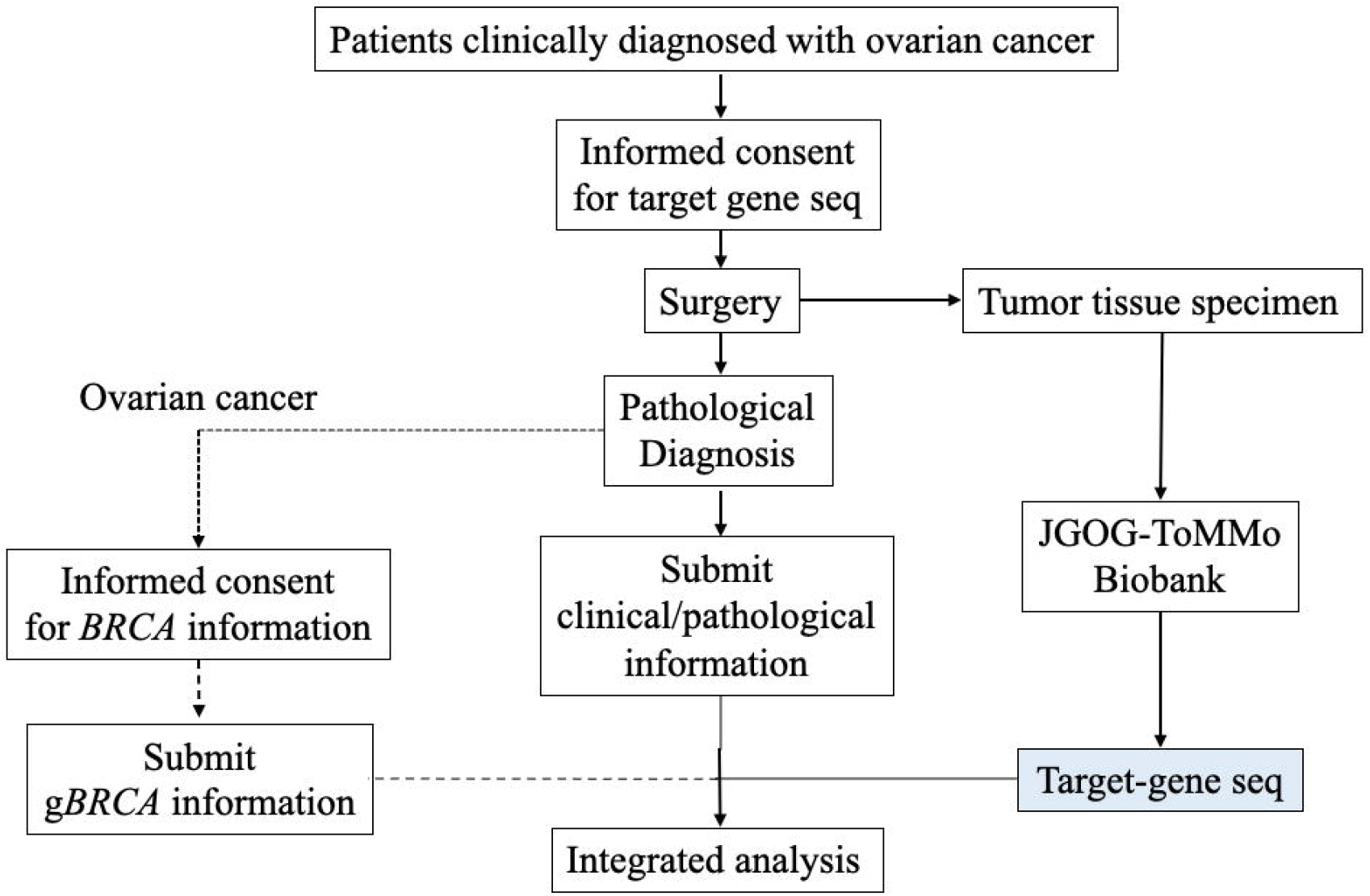
A schema of JGOG 3025 trial.

We collected data including patient characteristics, clinicopathological data, clinical outcome, and adverse events. Clinical outcome was updated prospectively every 6 months on the electronic data capture system.

The study was conducted in accordance with the Declaration of Helsinki and the Ethical Guidelines for Medical and Health Research Involving Human Subjects. Approval from the institutional review board of each participating JGOG institution was obtained prior to initiation of the study. All patients provided written informed consent.

### 2.2 Study population

The inclusion criteria are as follows: Patients who can approve informed consent and sign it; Patients who show their will to participate in this study and can sign the informed consent forms by themselves; Patients who are clinically diagnosed as ovarian cancer and can provide written informed consent before the surgery; Patients who are 20 years old and over at the enrollment; Patients with ECOG Performance status (PS) 0-2; Patients who can provide tumor tissue specimens. Ascites cytology and cell block specimens are not considered tumor tissue specimens. For patients to be treated with Neoadjuvant Chemotherapy (NAC), a tumor biopsy should be performed prior to NAC to make a pathologic diagnosis and to obtain a tumor tissue specimen.

Patients are excluded based on the following criteria: Patients with other clinically active malignancy (except breast cancer) that have been treated within 5 years (excluding basal cell carcinoma and squamous cell carcinoma of the skin, and carcinoma in situ or intramucosal carcinoma which are curable with local treatment); the principal investigator judges that enrollment of the patient in the study is inappropriate.

### 2.3 Study outcomes

The primary outcome of JGOG3025 trial is to identify the HRD frequency in Japanese patients with ovarian cancer including fallopian tube cancer and primary peritoneal cancer. The secondary outcome is to assess associations between PFS and HRD in ovarian cancer and between PFS and germline *BRCA1/2* mutation in ovarian cancer. PFS was defined as the time from registration in the trial to disease progression or death from any cause.

### 2.4 Definition of HRD

We performed target-gene sequencing of 51 genes by using DNA extracted from cancer tissues. A list of genes is shown in Table S2. Five genes (*ARID1A, KRAS, PIK3CA, PTEN*, and *TP53*) were included as molecular diagnostic markers in ovarian cancer, and 29 genes were categorized as HR-associated genes. We used QIAseq Targeted DNA Custom Panel (CDHS-16649Z-2451, Qiagen) and conducted 150bp paired-end sequencing on the Illumina NextSeq sequencer. Single nucleotide variant (SNV) or short InDel was detected by smCounter2 (Qiagen)^11^. Variant annotation was performed using ANOVAR version 20190408^12^.

To make a mutation list, we generated MAF file using maftools^13^ and filtered as follow: variants with variant allele frequency(VAF) less than 5% were excluded; variants classified as Nonsense, Frame Shift InDel, Nonstop, Traslation Start Site, Missense, Splice Site were included; variants with VAF more than 1% in gnomAD^14^, 1000 Genomes Project^15^, ExAC^16^, ESP6500^17^, or 14K Japanese custom reference from jMorp^18^ were excluded; variants interpreted as pathogenic or likely pathogenic in ClinVar (https://www.ncbi.nlm.nih.gov/clinvar/) or InterVar^19^ were included; Based on dbsnpf35a or dbscsnv11, Missense variants with deleterious 2 predictions or Splice variants with deleterious > 1 prediction were included; variants which are not found in both COSMIC^20^ and ICGC^21^ were excluded.

As for HRD, we defined case harboring at least one mutation of HR-associated genes as HRD and case without any HR-associated gene mutation as non-HRD. Because Microsatellite Instability (MSI)-high or POLE mutated ovarian cancers showing very high tumor mutational burden led to passenger mutation of HR-associated genes, cases with both HR associated gene mutations and MSI-high or POLE mutations were determined as HRD negative.

### 2.5 Central pathological review

To confirm the histological diagnosis, central pathological review was performed by three pathologists assigned by the Japanese Gynecologic Oncology Group using hematoxylin and eosin slide specimens from tumor tissues. The central histological evaluation and diagnosis of ovarian cancer was based on the WHO classification of tumors of female reproductive organs.

### 2.6 Statistical analysis

Descriptive statistics were used for clinicopathological characteristics, with n (%) for categorical variable. The full analysis set (FAS) consisted of all ovarian cancer patients with target-gene sequencing data. The frequency of HRD was calculated, together with 95% CIs, for all patients, and was stratified by FIGO stage and histologic types. Cox proportional hazard model analysis was performed to investigate an association of HRD or germline *BRCA1/2* status with PFS or OS after adjustment for age, stage, histologic type, surgery, NAC, and adjuvant chemotherapy. All statistical analyses were performed using SAS 9.4 (SAS Institute, Cary, North Carolina, USA) or R version 4.1.1 (https://www.R-project.org/.).

## 3. Results

### 3.1 Patient characteristics

The study was conducted from June 2015 to November 2019. The enrollment period was initially planned for 2 years but was extended to 2 years 3 months to reach the sample size required. The patient disposition is shown in Figure 2. A total of 996 patients who were clinically diagnosed with ovarian cancer were enrolled, but 295 patients were excluded from the full analysis set according to the eligibility criteria. A total of 701 patients were included in the final analysis. Patient characteristics are shown in Table 1. Patients in the full analysis set had a mean age of 58.4 years. Of 701 patients, 47.4% were stage I/II and 51.6% were stage III/IV.

**Figure 2.**
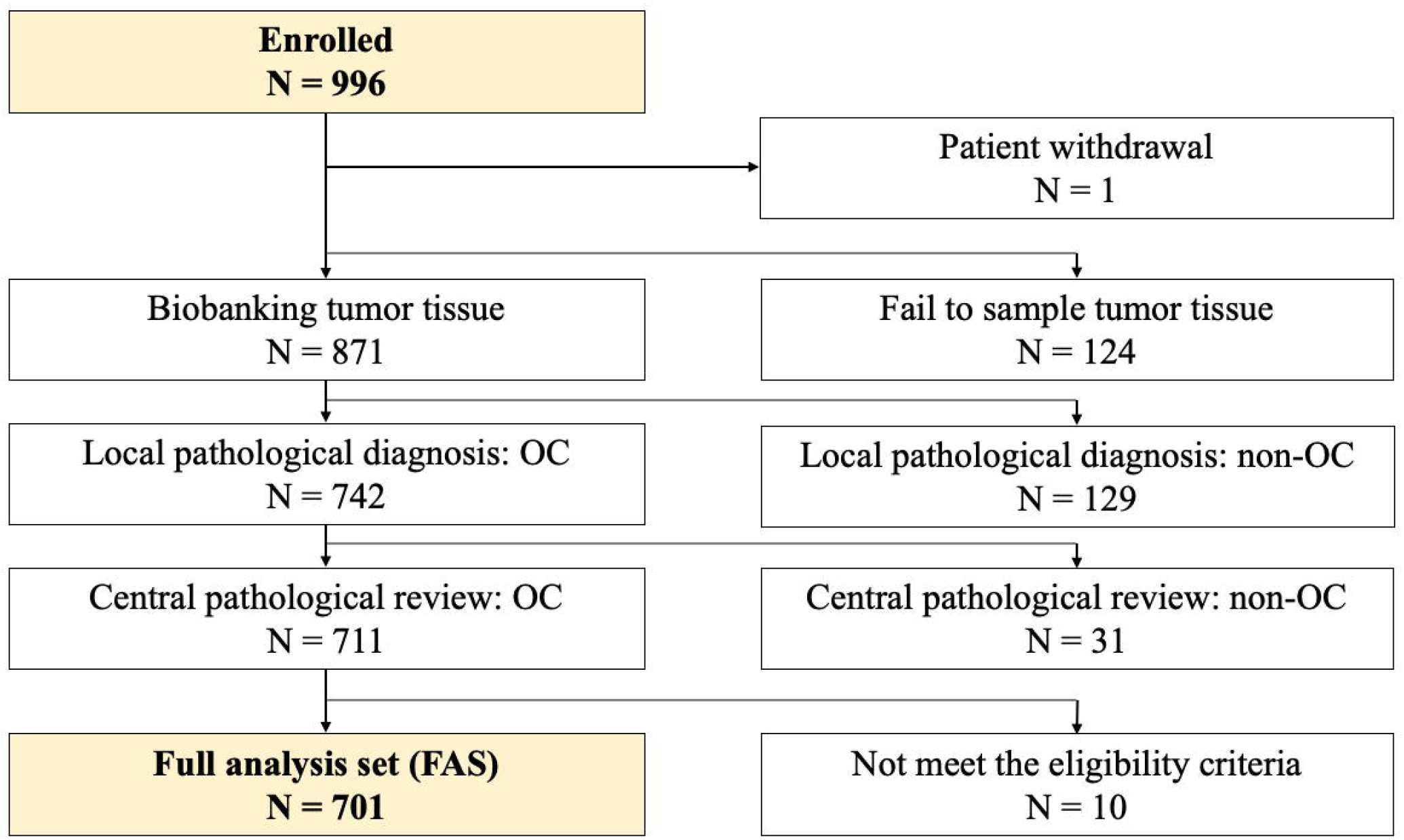
Patients disposition.

**Table 1.** Clinicopathological characteristics of FAS.

|  | FAS (n = 701) |
| --- | --- |
| Age |  |
| < 65 years | 474 (67.6%) |
| ≥ 65 years | 227 (32.4%) |
| FIGO stage |  |
| I/II | 332 (47.4%) |
| III/IV | 362 (51.6%) |
| Unknown | 7 (1.0%) |
| Histologic type |  |
| High-grade serous carcinoma | 298 (42.5%) |
| Clear cell carcinoma | 189 (27.0%) |
| Endometrioid carcinoma | 135 (19.3%) |
| Mucinous carcinoma | 12 (1.7%) |
| Low-grade serous carcinoma | 3 (0.4%) |
| Others | 64 (9.1%) |
| Surgery |  |
| Complete | 520 (74.2%) |
| Optimal (<1cm) | 75 (10.7%) |
| Not optimal (≥1cm) | 104 (14.8%) |
| Unknown | 2 (0.3%) |
| Neoadjuvant chemotherapy |  |
| Yes | 152 (21.7%) |
| No | 549 (78.3%) |
| Adjuvant chemotherapy |  |
| Yes | 612 (87.3%) |
| No | 89 (12.7%) |

### 3.2 Primary outcome

The frequency of HRD in Japanese patients with ovarian cancer was 45.2% (317/701) (Figure 1). The mutation rates of tumor *BRCA1* and *BRCA2* were 9.8%, 7.7%, respectively. Seven patients (1.0%) had both germline *BRCA1* and *BRCA2* mutations. On the other hand, the mutation rate of HR-associated genes except *BRCA1/2* was 26.7%.

### 3.3 Secondary outcomes

First, we performed Cox proportional hazard model analysis after adjustment for age, stage, histologic type, surgery, NAC, and adjuvant chemotherapy. There was no significant difference in PFS between HRD and non-HRD groups (adjusted hazard ratio (aHR), 0.84; 95% confidence interval (CI), 0.66-1.06; p = 0.14).

Next, we stratified ovarian cancers by FIGO stage. Although there was no significant difference in PFS between stage I/II HRD and non-HRD subgroups (aHR, 1.35; 95%CI, 0.82-2.24; p = 0.24), PFS in stage III/IV HRD subgroup was significantly longer than that in non-HRD subgroup (aHR, 0.72; 95%CI, 0.55-0.93; p = 0.013) (Figure 4). In addition, OS in stage III/IV HRD subgroup was also significantly longer than that in non-HRD subgroup (aHR, 0.58; 95%CI, 0.39-0.87; p = 0.009) (Figure 5).

**Figure 3.**
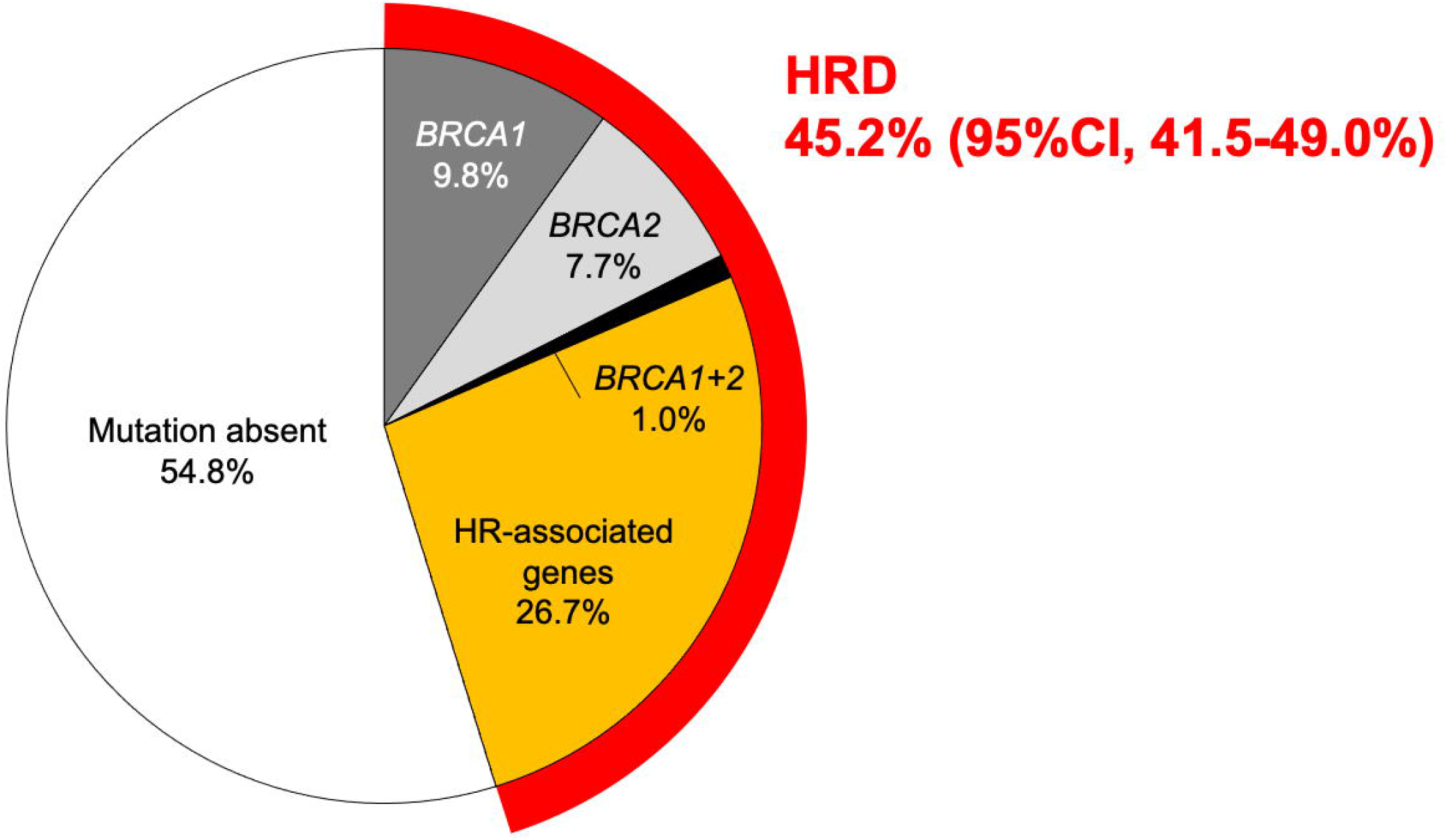
Frequency of HRD in Japanese patients with ovarian cancer.

**Figure 4.**
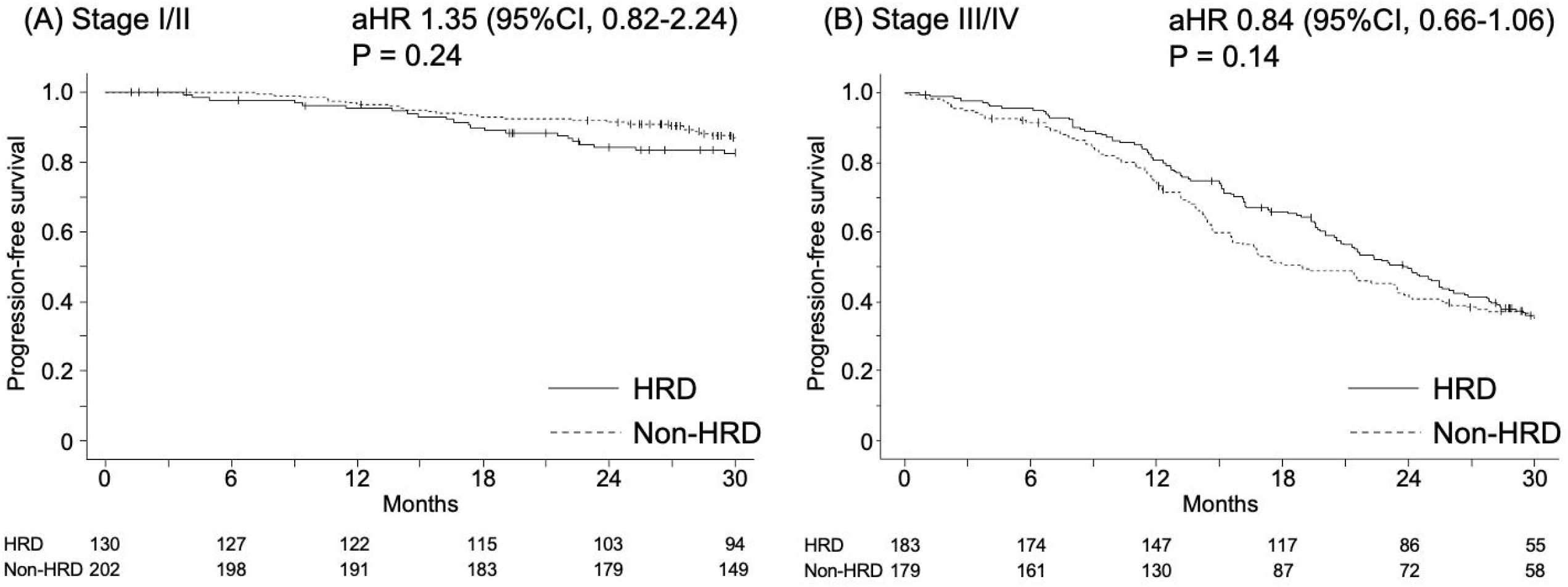
Association between PFS and HRD in Japanese patients with ovarian cancer.

**Figure 5.**
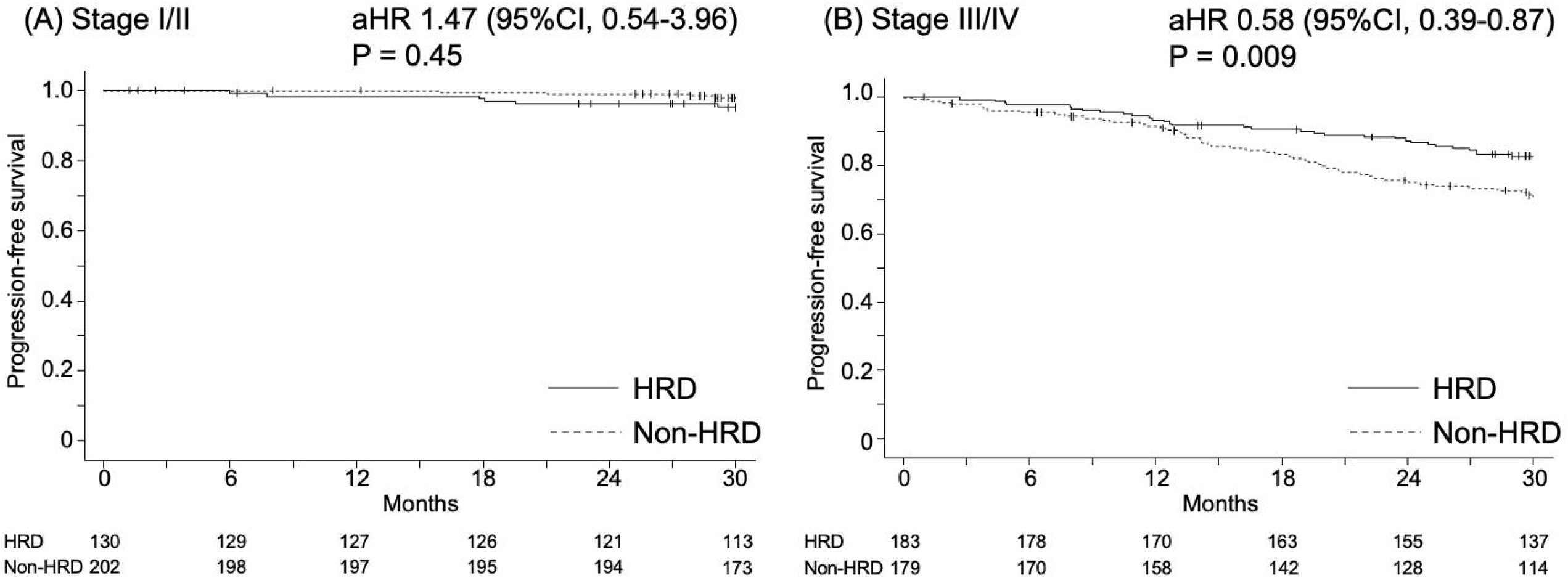
Association between OS and HRD in Japanese patients with ovarian cancer.

The frequency of HRD by histologic type was shown in Table 2. In patients with high-grade serous carcinoma, the frequencies of both HRD and tumor *BRCA1/2* mutations were relatively higher than those of other histologic types. Although the frequency of HRD in clear-cell type was also relatively high, tumor *BRCA1/2* mutation rate was lower than that of high-grade serous carcinoma.

**Table 2.** The frequency of HRD by histologic type.

| Histology | N | HRD | *BRCA1* | *BRCA2* | *BRCA1+2* |
| --- | --- | --- | --- | --- | --- |
| High-grade serous carcinoma | 298 | 154 (51.7%) | 52 (17.4%) | 33 (11.1%) | 2 (0.7%) |
| Clear cell carcinoma | 189 | 81 (42.9%) | 6 (3.2%) | 2 (1.1%) | 0 (0%) |
| Endometrioid carcinoma | 135 | 49 (36.3%) | 6 (4.4%) | 16 (11.9%) | 3 (2.2%) |
| Mucinous carcinoma | 12 | 3 (25.0%) | 0 (0%) | 0 (0%) | 0 (0%) |
| Low-grade serous carcinoma | 3 | 3 (100%) | 0 (0%) | 0 (0%) | 0 (0%) |
| Others | 64 | 27 (42.2%) | 5 (7.8%) | 3 (4.7%) | 2 (3.1%) |

Finally, we focused on germline *BRCA1/2* mutation status in ovarian cancer. We collected germline *BRCA1/2* information from 112 participants. Of them, germline *BRCA1/2* mutations were positive in 21 participants. 18 of which were identified in the corresponding tumors in this study cohort. There was no significant difference in PFS between PFS and germline *BRCA1/2* in stage I/II ovarian cancer because only three participants harbor germline *BRCA1/2* mutation (aHR, 1.12; 95%CI, 0.08-15.67; p = 0.93). On the other hand, germline *BRCA1/2* mutated stage III/IV ovarian cancers show significantly longer PFS compared to germline *BRCA1/2* wildtype stage III/IV ovarian cancers (aHR, 0.32; 95%CI, 0.14-0.72; p = 0.006) (Figure 6). In addition, germline *BRCA1/2* mutation was significantly associated with OS in stage III/IV ovarian cancer (aHR, 0.12; 95%CI, 0.02-0.59; p = 0.009).

**Figure 6.**
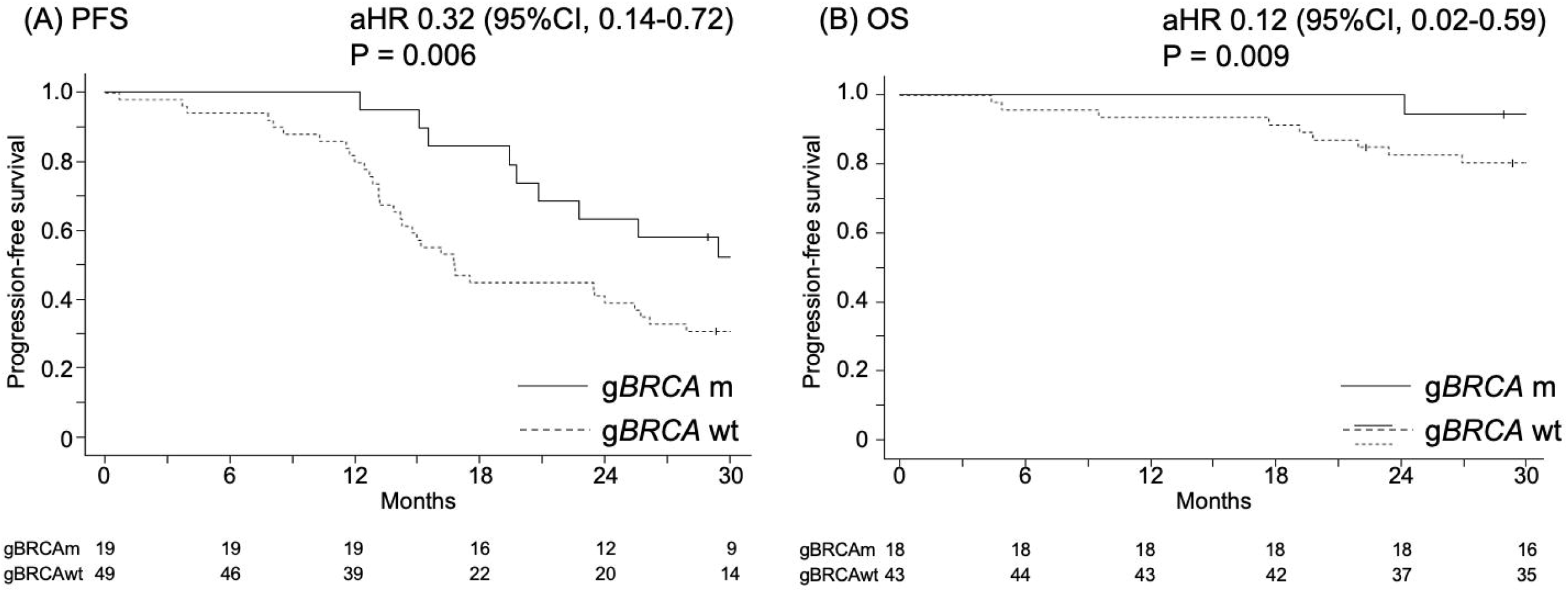
Association of germline *BRCA1/2* mutation with PFS and OS in stage III/IV ovarian cancer.

## 4. Discussion

Our multicenter collaborative prospective observational study revealed that the frequency of HRD in Japanese patients with ovarian cancer was 45.2% and that HRD was significantly associated with PFS and OS in advanced stage ovarian cancer.

In this study, we enrolled patients who were clinically diagnosed with ovarian cancer and could provide tumor tissue sample after surgery. Compared to the real-world distribution of each histology based on the JSOG annual report^22^, the proportion of mucinous carcinoma and low-grade serous carcinoma was smaller in the JGOG3025 trial. This may be due to the difficulty of preoperative diagnosis and tumor tissue sampling in mucinous and low-grade serous carcinomas. HRD was significantly associated with prognosis in Stage III/IV ovarian cancer but not in Stage I/II. The distribution of histologic types differed between Stage I/II and Stage III/IV, and the proportion of high-grade serous carcinoma was higher in Stage III/IV than in Stage I/II. Therefore, the frequency of *BRCA1/2* mutations was also higher in Stage IIII/IV. It is well known that *BRCA1/2* mutation is associated with platinum sensitivity in ovarian cancer^23^, and *BRCA1/2* status might affect the present results. On the other hand, we also found an association between PFS and HRD in Stage III/IV clear cell carcinoma whish harbored less *BRCA1/2* mutation. These results suggest that not only *BRCA1/2* but also other HR-associated gene mutations are linked with platinum sensitivity in advanced-stage ovarian cancer^24^.

Olaparib maintenance therapy for platinum-sensitive relapsed ovarian cancer was approved by health insurance in January 2018 in Japan. The SOLO2 trial has shown that Olaparib treatment group has longer OS (median 51.7 months; 95%CI 41.5-59.1) than the placebo group (median 38.8 months; 95%CI 31.4-48.6) (HR 0.74; 95%CI 0.54-1.00; p = 0.054) although 38% of the placebo group received subsequent PARP inhibitor therapy^25^. In addition, niraparib for platinum-sensitive relapsed or HRD ovarian cancers was also approved by health insurance in September 2020. Therefore, it is necessary to verify whether there is a difference in the rate of PARP inhibitor administration between HRD and non-HRD groups.

In this study, HRD was defined as the presence of at least one HR-related gene mutations. In clinical practice, Myriad myChoice diagnostic system is now covered by health insurance in Japan and is the standard method for assessing genomic instability (GI) score calculated by genomic loss of heterozygosity (LOH)^26^, telomeric allelic imbalance (TAI)^27^, and large-scale state transition (LST)^28^. Recently, the assessment of HRD based on mutational signature is also proposed^29, 30^. However, the two methods reflect the genomic scar caused HRD and do not indicate whether the current tumor is HRD or not. On the other hand, HR-associated gene mutation is one of the etiologies of HRD. It is well known that *BRCA1/2* mutations are significantly correlated with GI score and significantly associated with clinical outcome in ovarian cancer. However, the clinical significance of HR-associated genes except *BRCA1/2* remains unclear. Recently, PAOLA-1 subsequent analysis demonstrated that HR-associated gene mutation (excluding *BRCA1/2* mutation) was not predictive of PFS benefit with maintenance olaparib in combination with bevacizumab, compared with bevacizumab alone, in PAOLA-1 trial and that only five genes (*BLM, BRIP1, RAD51C, PALB2*, and *RAD51D*) had a median GI score ≥ 42. Further multi-omics data analysis should be needed to the clinical impact of each HR-associated gene mutation in ovarian cancer.

Our study has several limitations. The HR-associated mutations may be monoallelic, without any alterations in the opposite allele. Although tumors with MSI-High or POLE mutations were excluded from HRD, other HR-associated mutations may also not contribute to tumorigenesis.

As correlation between GI score and HR-associated mutations has not been analyzed, the HRD status in each tumor has not been fully verified. In addition, the target sequencing cannot pick up tumors with epigenetic alterations (such as hypermethylation in promoter region in *BRCA1* or *RAD51C*).

In conclusion, HRD was detected in not only high-grade serous histologic type but also other histologic types, and HRD status may be useful to provide an effective treatment option for patients with ovarian cancer.

## Data Availability

All data produced in the present study are available upon reasonable request to the authors.

## Abbreviations

aHR: adjusted hazard ratio
CI: Confidence Interval
FAS: Full analysis set
FIGO: International Federation of Gynecology and Obstetrics
GI: Genomic Instability
HR: homologous recombination
HRD: homologous recombination deficiency
JGOG: Japanese Gynecologic Oncology Group
MSI: Microsatellite instability
NAC: Neoadjuvant chemotherapy
OS: Overall survival
PARP: poly (ADP-ribose) polymerase
PFS: Progression free survival
PS: Performance status
SNV: Single nucleotide variant
TCGA: The Cancer Genome Atlas
ToMMo: Tohoku Medical Megabank organization
VAF: variant allele frequency

## Acknowledgments

The authors wish to thank all members of the Japanese Gynecologic Oncology Group. We appreciate all women who participated in this study and their families, the staff of Translational Research Center for Medical Innovation (TRI; the data and statistical analysis center for JGOG3025), and the participating JGOG member institutions.

## Ethics Statement

1. Cohort name: Japanese patients with ovarian cancer cohort (JGOG3025)
2. Non-abbreviated, full name of Ethics Committee / Institutional Review Board (IRB) for the cohort: The Ethics Committee of the Japanese Gynecologic Oncology Group and Niigata University
3. Decision made by ethics oversight body: approved (No. JGOG3025 in the Ethics Committee of JGOG and No. 2006-0006 in Niigata University)
4. Informed Consent: All patients provided written informed consent.
5. Registry and the Registration No. of the study/trial: NCT03159572 (ClinicalTrials.gov); UMIN 000026303 (UMIN-CTR)
6. Animal Studies: N/A

